# Assessment of Medication Adherence in Patients: Development and Validation of a Machine Learning Model

**DOI:** 10.1101/2025.09.14.25335382

**Authors:** Jifan Zhang, Jason Zhensheng Qu

## Abstract

**Background:** This study addresses limitations of traditional medication adherence assessment tools by developing a machine learning model to evaluate post-discharge medication compliance in patients using drugs. The research was conducted at Nanjing Drum Tower Hospital from February 2024 to December 2024.

**Methods:** We collected clinical data from 240 patients through questionnaires and developed a multi-class machine learning model. Feature selection employed manual screening and polynomial logistic regression. Six ML models were evaluated, with the Random Forest Classifier (RFC) demonstrating optimal performance (bad_AUC = 0.979, fine_AUC = 0.973, good_AUC = 0.917). SHAP analysis was used to explain the best-performing model.

**Results:** The RFC model showed superior predictive capability across all adherence levels. Model interpretation revealed key clinical factors influencing adherence patterns. The tool enables early identification of non-compliance and supports intervention strategies.

**Conclusions:** This RFC-based model represents a significant advancement in medication adherence assessment, offering clinicians a practical tool for monitoring compliance. The approach shows particular promise for enhancing mental health management in this patient population by fostering better medication awareness and establishing scientific medication habits during early treatment stages.

---

As a core determinant of treatment efficacy, medication adherence has become a significant research topic in the field of precision medicine [1]. The World Health Organization (WHO) has highlighted that treatment failures, complications, and increased healthcare costs caused by long-term non-adherence to medication regimens have evolved into major societal issues [2]. Medication adherence is influenced by multi-dimensional factors: patient age, health literacy, and psychological state form internal drivers; medication dosage frequency and side effects create direct barriers; while the quality of medical communication and socioeconomic conditions constitute external constraints [3]. However, existing studies primarily focus on specific disease populations, lacking systematic attention to medication behaviors in the general population, which fails to meet the needs of the precision medicine era. Current mainstream medication adherence assessment tools (e.g. MMAS-8, MARS scale) have significant limitations [4]. The MMAS-8 scale measures patients’ medication-taking behavior over the past four weeks through 8 items, but patients may overestimate adherence due to social desirability bias, while clinical records often overlook asymptomatic medication interruptions [5-6]. Traditional scales rely on patient self-reports or clinical observations, making them susceptible to recall bias and subjective judgments, and most are static assessments unable to capture dynamic changes in medication behaviors [7-8]. This study innovatively constructs a predictive model: using machine learning algorithms to identify high-risk non-adherent populations, combined with other technologies to implement precision interventions, ultimately achieving full-cycle management of treatment.

## Materials and methods

### Study design

The study protocol strictly follows the Transparent Reporting of a Multivariable Prediction Model for Individual Prognosis or Diagnosis (TRIPOD) reporting guidelines.

#### Participants

This study was conducted from February 2024 to December 2024 in the inpatient department of Nanjing Drum Tower Hospital. All subjects in this study were patients who had used drugs. The inclusion criteria were: (1) Chinese citizens aged ≥ 18 years, (2)voluntary participation in the survey and (3) being able to complete the related questionnaires independently. The exclusion criteria were: (1) prior medical history with severe mental illnesses, (2) prior use of medical drugs to treat mental disorders, (3)pregnant and lactating women, (4) medical history of malignant tumor.

#### Data collection

We collected comprehensive patient data in a structured format, including demographics (age, height, weight, gender, marital status, educational attainment, type of medical insurance), comorbidity history (including hypertension, diabetes, coronary heart disease, gout, renal insufficiency, liver dysfunction, pulmonary hypertension, heart failure), medication use (aspirin, clopidogrel, amiodarone, NASIDs (including tramadol), digoxin, beraprost, ACEI/ARB, β-blockers, statins), laboratory test results (TTR, variability classification, regular follow-up, location, number of comorbid diseases, number of concomitant medications, polypharmacy, disability, multiple diseases, history of thrombosis, history of stroke, history of bleeding, whether taking medication), and other relevant metrics. For the assessment of medication adherence, we utilized a well-validated and reliable tool to measure the adherence behavior of patients over the past two weeks. Medication adherence was categorized based on standard criteria: bad adherence, fine adherence, and good adherence. In this study, we formulated medication adherence prediction as a three-class classification task: bad adherence, fine adherence, and good adherence.

#### Data preprocessing

We performed min–max normalization on the entire dataset by mapping all features to the range of [0, 1] using the maximum and minimum values of each feature. This normalization eliminates scale differences among different features and ensures comparability and analysis within the same numerical range, thereby enhancing the performance of the algorithm model.In machine learning(ML) modeling, a sample class ratio lower than 9:1 is typically regarded as imbalanced. Our dataset had a notably lower proportion for one outcome event, indicating an extremely skewed class distribution. To address this, we used the Synthetic Minority Over - sampling Technique (SMOTE) to generate a balanced dataset. This approach involves synthesizing samples for the minority class to balance the number of samples across different classes, thus creating a dataset where each class has a similar representation for more effective modeling.

#### Feature selection

A two-step feature selection process was employed. First, manual screening identified potentially relevant features associated with medication adherence. Subsequently, polynomial logistic regression was applied to further refine the feature subset. This method helped in selecting the most significant features that could potentially influence medication adherence.

#### Data imputation and normalization

Missing values were imputed using K-Nearest Neighbors (KNN) for continuous variables and mode imputation for categorical variables in both the training and test sets. Additionally, continuous features were normalized using Z-score standardization, while multi-categorical features were one-hot encoded to create dummy variables, prior to model training.

#### Model development

A total of 240 patients were involved in this study.The normality of continuous variables was evaluated using the Shapiro-Wilk test. For normally distributed variables, the mean and standard deviation were utilized to express their values, whereas non-normally distributed variables were represented by the median (interquartile range [IQR]). When comparing two groups, Student’s t-test or the Wilcoxon rank-sum test was selected based on the distribution characteristics of the data. Categorical variables were presented in terms of counts and percentages, with comparisons between groups conducted using the chi-square test. Variables with more than 10% missing values were excluded from the analysis. The remaining missing values were imputed using a random forest-based multiple imputation method. To address the class imbalance between the negative and positive groups, the Synthetic Minority Over-sampling Technique (SMOTE) was applied to balance the dataset. Finally, the dataset was divided into training and validation sets at a ratio of 80% to 20%.Grid search was employed for hyperparameter tuning. For variable selection, classical polynomial logistic regression was utilized. Variables with a p - value less than 0.05 were included in the analysis, while the rest were excluded.

To enhance the reliability and generalization ability of the model, a five - fold cross - validation was implemented. In this process, the training set was further divided into five non - overlapping subsets. In each iteration, four subsets were used for training, and the remaining one subset was used for validation. This approach was repeated five times, with each subset serving as the validation set exactly once. By averaging the evaluation metrics across these five iterations, a more comprehensive and accurate assessment of the model performance was obtained.

Six machine learning models were adopted, namely logistic regression, support vector machine, K - Nearest Neighbors (KNN), Random Forest Classifier (RFC), CatBoost, and LightGBM (LBGM). These models were trained and evaluated to predict the anticoagulation outcomes, aiming to establish a reliable prediction model for better understanding and predicting the efficacy of anticoagulation therapy.

#### Model evaluation

The hold-out test set was used exclusively to evaluate the generalizability of the developed models. Considering the inherent characteristics of the multi-classification task, we employed a comprehensive suite of evaluation metrics to assess multi-classification model performance. This included: (1) Confusion Matrix: Visualized the distribution of true and predicted medication adherence classifications across categories (bad, fine, good); (2) Multi-class ROC Curves: Illustrated the tradeoff between sensitivity and specificity for each adherence class (see Fig. 2 for the ROC curves of the six models); (3) Macro-average AUC: Evaluated overall model performance by averaging AUC scores across all adherence categories; (4) Kappa Statistic: Assessed model agreement with the true classifications, accounting for class imbalance; (5) Average Precision, Recall, and F1-score. In a clinical setting, the ideal approach would be to develop a predictive model that is more effective at identifying patients with poor medication adherence.

**Fig. 1.**
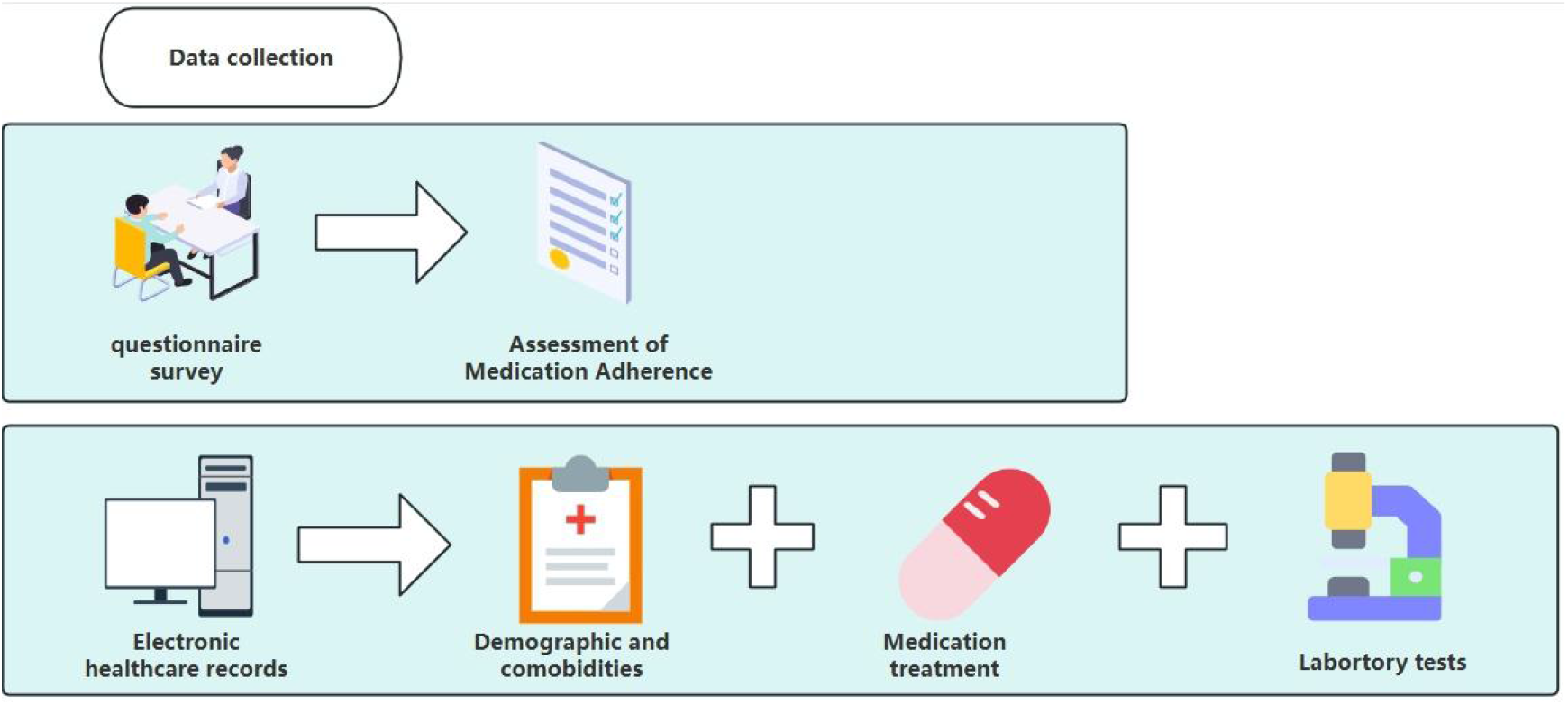
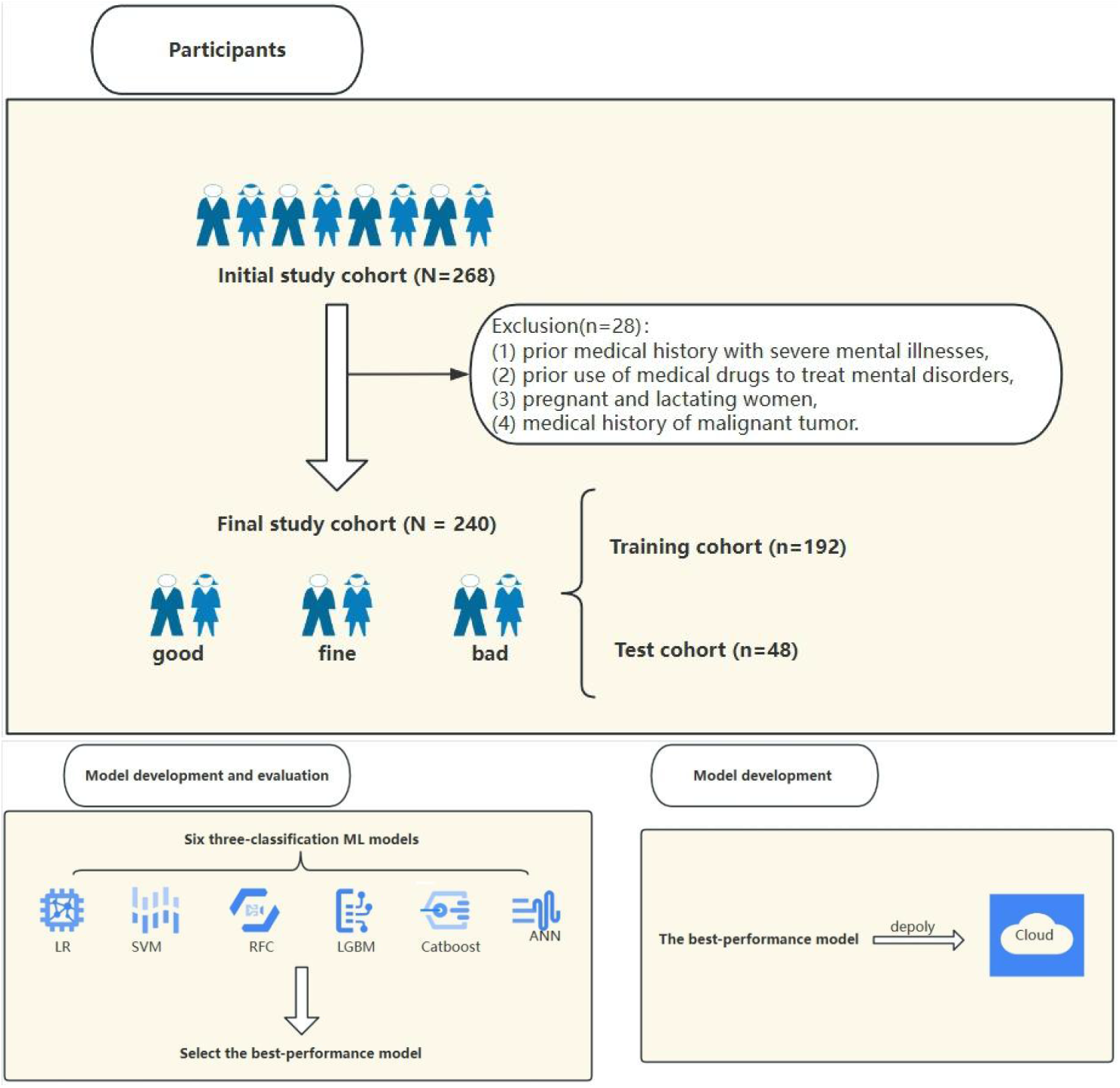
Overview of the study workflow.

**Fig. 2.**
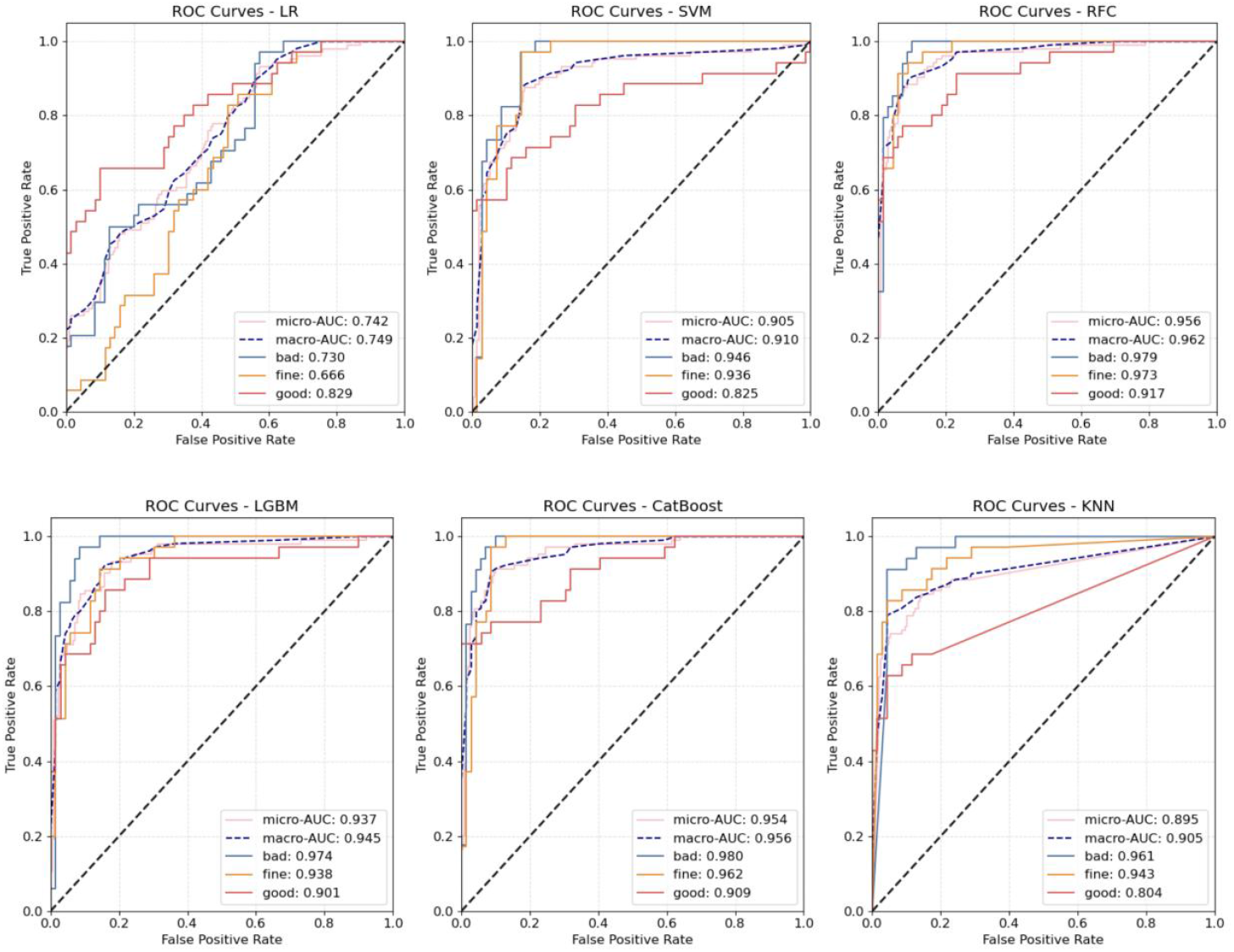
ROC curve of the six multi-classification ML models in the test cohort. (a) LR; (b) SVM; (c) RFC; (d) LGBM; (e) Catboost; (f) ANN. Notes: ROC, receiver operating curve; ML, machine learning; LR, logistic regression; SVM, support vector machine; RFC, random forest classifier; LGBM, light gradient boosting machine; Catboost, categorical boosting; ANN, artificial neural network.

Furthermore, the validity of the proposed models was assessed using the test cohort. The performance of the six models within the test cohort is detailed in Table 1 and illustrated in Fig. 2. Confusion matrix of the six multi-classification ML models in the test cohort is shown in Fig. 3. Among these, RFC model demonstrating strong predictive capabilities across different levels of medication adherence (bad_AUC = 0.979412, fine_AUC = 0.973499, good_AUC = 0.917184). These results highlight the robust performance of the RFC model.

**Table 1.** The performance of the following six multi-classification ML models in the test cohort (n=48). ML, machine learning; LR, logistic regression; SVM, support vector machine; LGBM, light gradient boosting machine; Catboost, categorical boosting; ANN, artificial neural network.

|  | LR | SVM | RFC | LGBM | CatBoost | KNN |
| --- | --- | --- | --- | --- | --- | --- |
| Accuracy | 0.528846 | 0.769231 | 0.865385 | 0.836538 | 0.865385 | 0.788462 |
| Kappa | 0.292812 | 0.654054 | 0.798142 | 0.754955 | 0.798170 | 0.683015 |
| Weighted F1 | 0.523127 | 0.767144 | 0.863920 | 0.831792 | 0.862914 | 0.775166 |
| Micro AUC | 0.742234 | 0.904863 | 0.956130 | 0.937454 | 0.954466 | 0.894693 |
| Bad_AUC | 0.730252 | 0.946218 | 0.979412 | 0.973529 | 0.979832 | 0.960924 |
| Fine_AUC | 0.666253 | 0.936232 | 0.973499 | 0.938302 | 0.961905 | 0.942650 |
| Good_AUC | 0.828571 | 0.825259 | 0.917184 | 0.901035 | 0.909317 | 0.803520 |

**Table 2.** The demographic, clinical and laboratory characteristics of the different depression levels groups in the training cohort.

|  | **Total**<br>**(n = 519)** | **Bad**<br>**(n = 173)** | **Fine**<br>**(n = 173)** | **Good**<br>**(n = 173)** | **P** |
| --- | --- | --- | --- | --- | --- |
| Age (year) | 61.45 [55.0, 68.44] | 63.08 [58.88, 68.26] | 66.01 [56.05, 70.96] | 53.50 [41.25, 63.00] | < 0.0001* |
| Height(cm) | 164.71 [160.00, 169.00] | 164.10 [160.68, 165.02] | 164.77 [158.73, 170.00] | 167.00 [160.00, 174.00] | 0.0011* |
| Weight(kg) | 63.00 [55.82, 70.18] | 57.51 [53.40, 70.44] | 64.51 [59.16, 69.93] | 62.00 [55.00, 71.00] | 0.0753 |
| Number of comorbid diseases | 2.00 [1.00, 2.35] | 2.00 [1.67, 2.94] | 2.00 [1.00, 2.76] | 1.00 [1.00, 2.00] | < 0.0001* |
| Number of concomitant medications | 1.00 [1.00, 2.00] | 2.00 [1.00, 3.00] | 1.00 [1.00, 2.00] | 1.00 [1.00, 2.00] | < 0.0001* |
| Gender |  |  |  |  | 0.0004* |
| Female | 245 (59.0%) | 99 (71.2%) | 80 (58.0%) | 66 (47.8%) |  |
| Male | 170 (41.0%) | 40 (28.8%) | 58 (42.0%) | 72 (52.2%) |  |
| Location |  |  |  |  | < 0.0001* |
| Town | 243 (58.6%) | 115 (82.7%) | 104 (75.4%) | 24 (17.4%) |  |
| City | 172 (41.4%) | 24 (17.3%) | 34 (24.6%) | 114 (82.6%) |  |
| Educational attainment |  |  |  |  | < 0.0001* |
| Low | 224 (54.0%) | 108 (77.7%) | 84 (60.9%) | 32 (23.2%) |  |
| Middle | 152 (36.6%) | 30 (21.6%) | 50 (36.2%) | 72 (52.2%) |  |
| High | 39 (9.4%) | 1 (0.7%) | 4 (2.9%) | 34 (24.6%) |  |
| Hhistory of troke |  |  |  |  | 0.0354* |
| No | 385 (92.8%) | 128 (92.1%) | 134 (97.1%) | 123 (89.1%) |  |
| Yes | 30 (7.2%) | 11 (7.9%) | 4 (2.9%) | 15 (10.9%) |  |

**Fig. 3.**
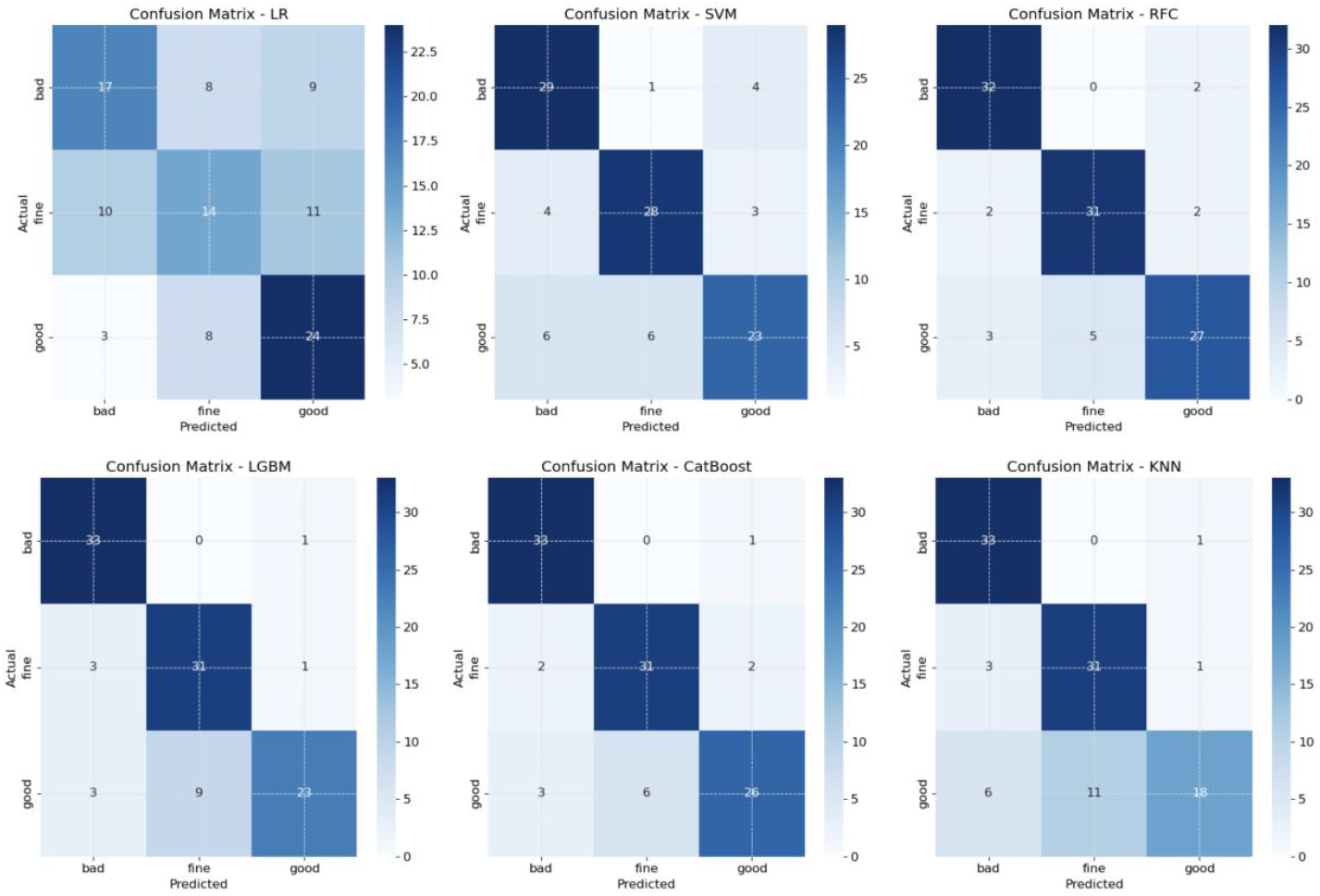
Confusion matrix of the six multi-classification ML models in the test cohort. (a) LR; (b) SVM; (c) RFC; (d) LGBM; (e) Catboost; (f) ANN. Notes: ML, machine learning; LR, logistic regression; SVM, support vector machine; RFC, random forest classifier; LGBM, light gradient boosting machine; Catboost, categorical boosting; ANN, artificial neural network.

#### Model interpretation

The SHapley Additive exPlanations (SHAP) technique, demonstrating each variable’s influence on the overall model, was further applied to gain insight into the best-performance model. We utilized the SHAP technique to evaluate the contribution of features in the best-performing model to the prediction outcomes of the test set. In the three-class classification task, the model’s objective is to accurately assign samples to one of the three categories. Consequently, we calculated the SHAP values for each category. For each sample, the SHAP value is represented as a matrix of size (1, k, 3), where each matrix element corresponds to the contribution of a feature to a specific category (with k representing the number of features in the model). We generated separate SHAP bar and dot plots for each category using the test set data (see Fig.4 for an example of SHAP plots). Furthermore, to visualize the distribution of key features across different adherence levels in a sample of patients, we generated a grouped-clustered heatmap for ten randomly selected patients using the “pheatmap” package in R (version 4.2.1). Finally, for enhanced clinical usability, the best-performance multi-classification model was deployed as an R Shiny application, facilitating user-friendly model interaction.

### Statistical analysis

This study aimed to compare the differences in demographic, clinical, and laboratory characteristics across three levels of medication adherence. The initial step involved evaluating the normality of continuous variables utilizing the Shapiro-Wilk test. Continuous variables that adhered to a normal distribution were presented as mean ± standard deviation (SD) and were analyzed using the Chi-square test. In contrast, those not following normal distribution were depicted as median ± interquartile range (IQR) and assessed with the Kruskal-Wallis test. Subsequently, categorical variables were compared using either the Chi-square (χ2) or Fisher’s exact test, depending on their distribution and sample size. A p-value of less than 0.05 was considered statistically significant for all tests. The entire statistical analysis process was performed using the “compareGroups” package in R version 4.2.1.

## Results

### Participants’ characteristics

The Morisky Medication Adherence Scale was used to assess the patients’ medication adherence. By filling out the questionnaires distributed by the hospital, a total of 240 eligible patients were included.The median age of the participants was 60.5 years (IQR: 51.2–69.8 years), with a predominance of female patients, accounting for 75.2%.To address the class imbalance between the negative and positive groups, the Synthetic Minority Over-sampling Technique (SMOTE) was applied to balance the dataset. Finally, the dataset was divided into training and validation sets at a ratio of 80% to 20%. These 240 patients were randomly divided into two groups: the training cohort (n = 192) and the test cohort (n = 48). Comparative analysis of baseline characteristics between the two cohorts indicated a general balance, as detailed in Supplementary Table 2.

The MMAS-8 scale was used to assess the medication adherence of patients. The medication adherence of patients was classified into three levels: bad adherence (below 6 points), fine adherence (6-8 points), and good adherence (8 points). Within the training cohort, the prevalence rates for these categories were 33.3% (n = 64/192), 33.3% (n = 64/192), and 33.3% (n = 64/192), respectively. Correspondingly, in the test cohort, the prevalence rates were 33.3% (n = 16/48), 33.3% (n = 16/48), and 33.3% (n = 16/48), respectively.

### Feature selection

In this research, forty-eight potential variables related to medication adherence in patients using drugs were initially considered. Of these, twenty-four variables were identified as having missing data. Twenty-four variables had missing values, and six variables with missing values over 25% were excluded from the analysis . Manual screening subsequently pinpointed fifteen variables markedly associated with the medication adherence outcome, including age, height, weight, BMI, number of comorbid diseases, number of concomitant medications, polypharmacy, disability, gender, regular follow-up, location, educational attainment, type of medical insurance, disease onset, and history of stroke.

These identified fifteen variables were further analyzed using polynomial logistic regression to discern the optimal subset. The polynomial logistic regression ultimately determined that ten variables had a significant association with the multi-classification medication adherence outcome: age, height, weight, number of comorbid diseases, number of concomitant medications, gender, location, educational attainment, history of stroke, and whether taking medication.

### Model performance

The aforementioned ten variables were used to construct the following six different multi-classification ML models: LR, SVM, RFC, LGBM, Catboost, and KNN. The optimal hyperparameters for each model are documented in Supplementary Table 3. Supplementary Table 4 and Figure 2 show the performance metrics of these models within the training cohort. Figure 3 shows the confusion matrix of each multi-class model on the training cohort. Notably, except for the LR and the KNN models, all other models demonstrated superior performance, ranking in the top four across various metrics, including bad_AUC, fine_AUC, and good_AUC. These models have the potential to correctly detect more positive cases of poor medication adherence.

**Table 3.**
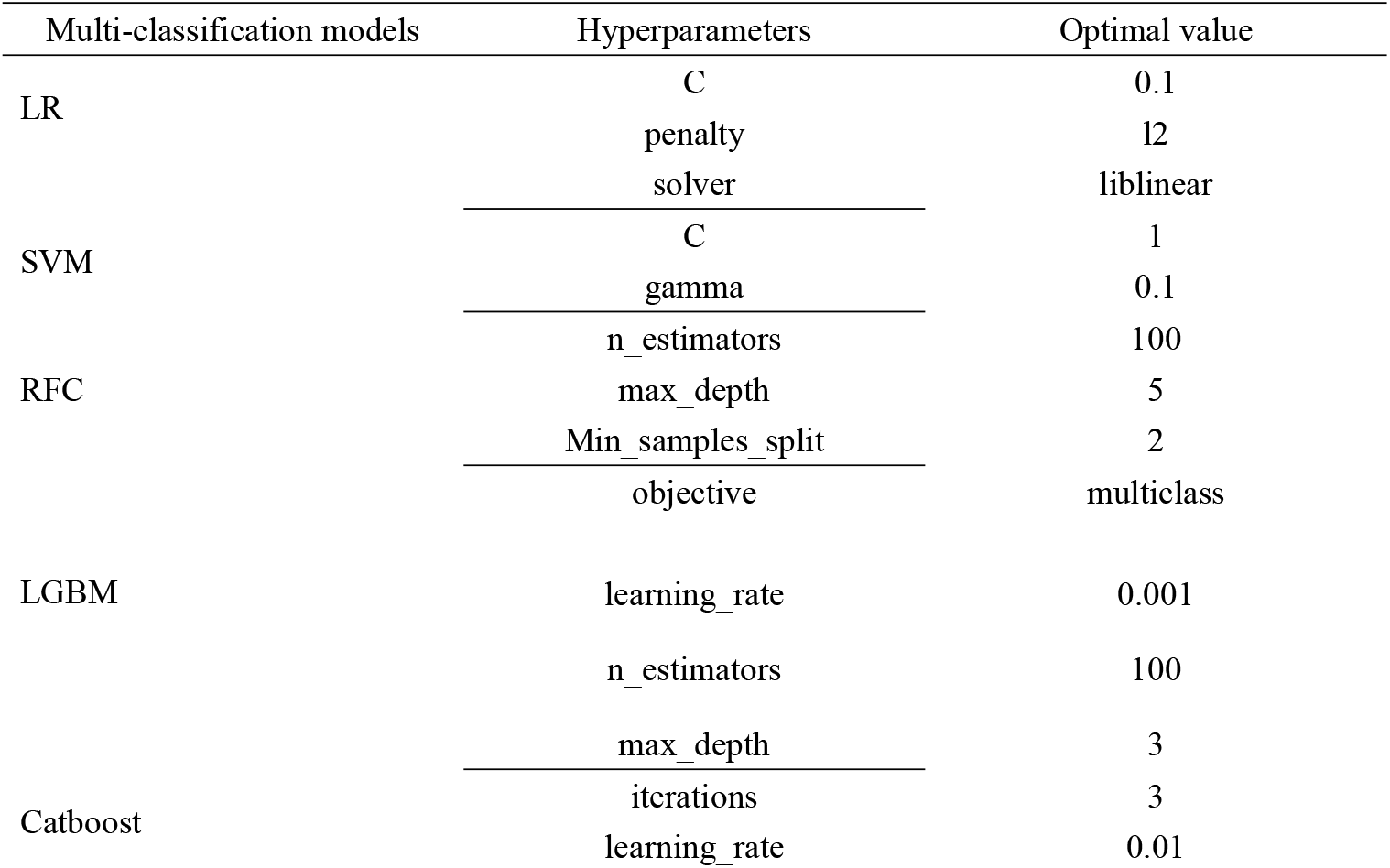
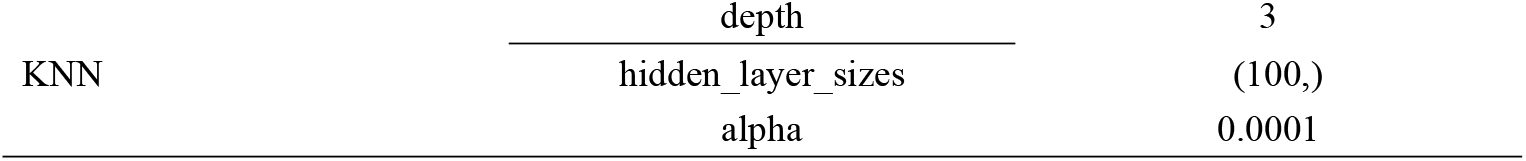
The best hyperparameters for the six multi-classification ML models.

**Table 4.**
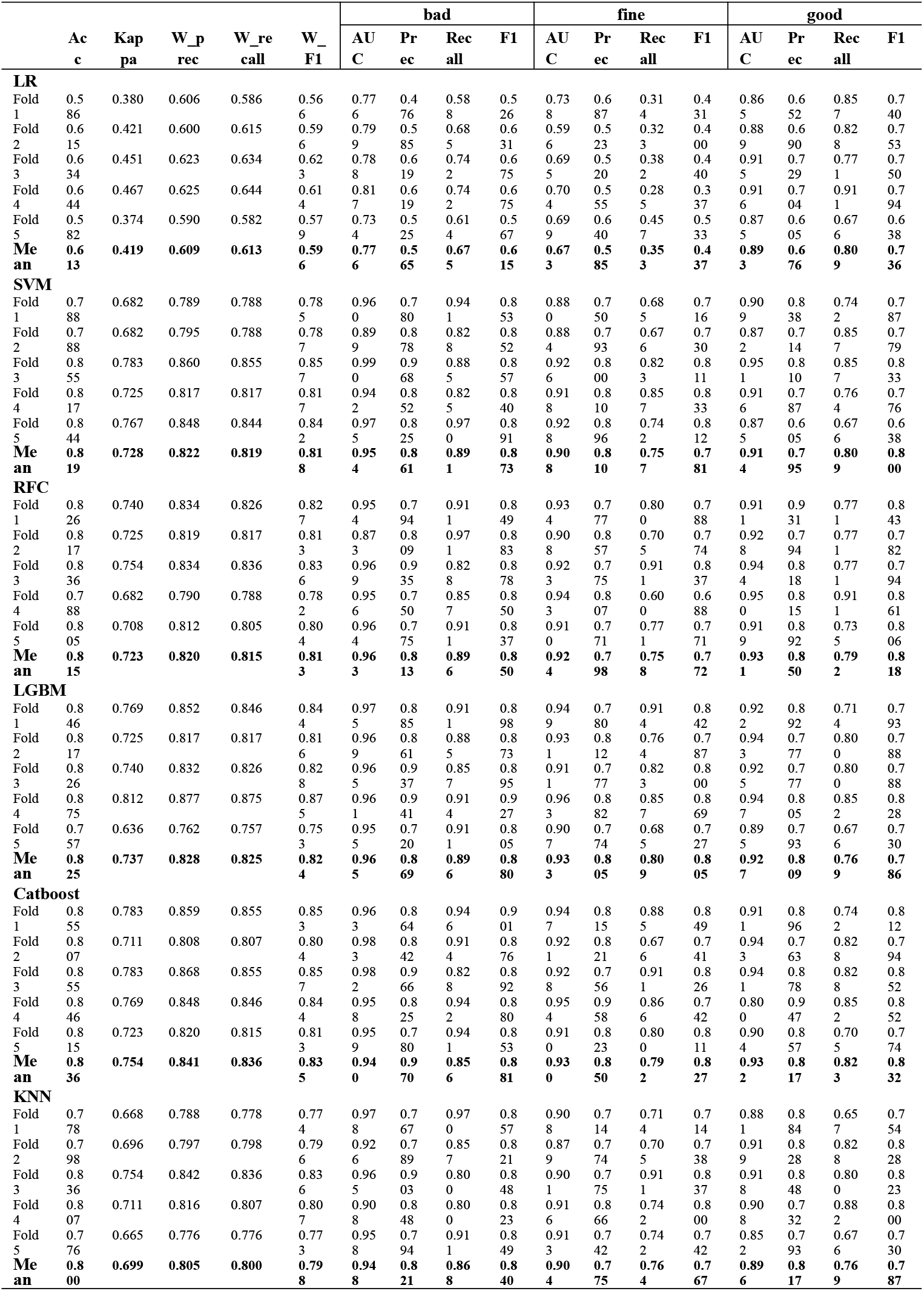
The performance of the six multi-classification ML models with 15-fold cross validation in the training cohort (n = 400)

Furthermore, the validity of the proposed models was assessed using the test cohort. The performance of the six models within the test cohort is detailed in Table 1 and illustrated in Fig. 2. Confusion matrix of the six multi-classification ML models in the test cohort is shown in Fig. 3. Among these, the RFC model exhibited the highest levels of bad_AUC, fine_AUC, and good_AUC. These results highlight the robust performance of the RFC model and its effectiveness in classifying medication adherence levels in patients using anticoagulant drugs.

### Model interpretation

The RFC model was explained through the application of the SHAP algorithm, which calculated the average absolute SHAP values to quantify the impact of each variable. Figure 4 illustrates the distributions of feature importance analyses and variable impact model outputs for the RFC model across the bad, fine, and good adherence categories. Notably, the analysis highlights that higher age, lower height, higher weight, increased number of comorbid diseases, higher number of concomitant medications, male gender, urban location, lower educational attainment, and history of stroke were associated with increased SHAP values. This suggests these factors correlate with an elevated risk of poor medication adherence, as indicated by the model.

**Fig. 4.**
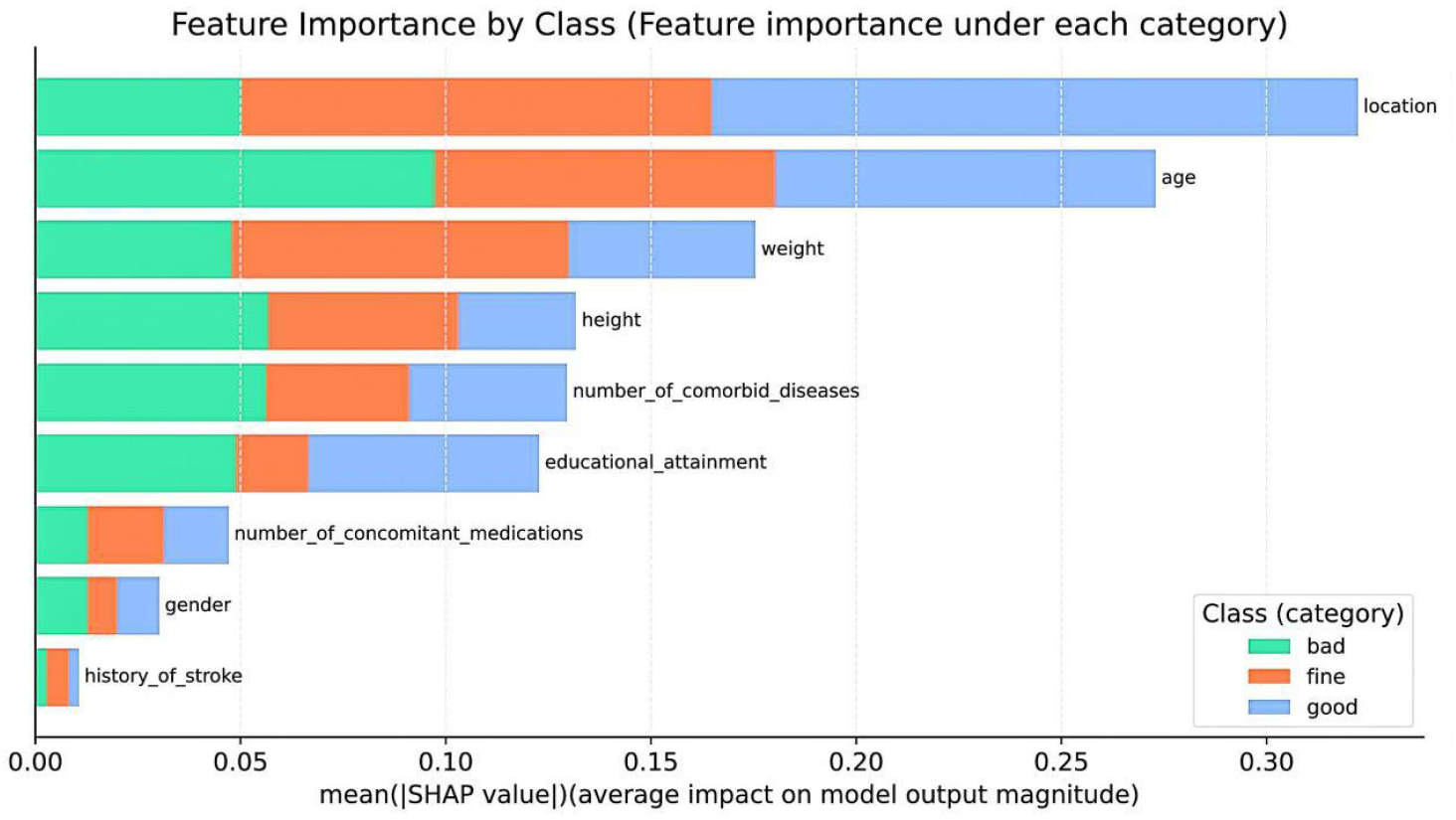
SHAP summary plots for explaining the nine variables contributing to the RFC model.

### Model deployment

To enhance the practical applicability of our proposed RFC model in clinical settings, the model was deployed to the cloud, making it accessible to healthcare professionals for real-time medication adherence assessment in patients using drugs. The model is hosted on a web-based platform available at https://patient-medication-adherence-calculator.streamlit.app/.

The user interface of this online tool is shown in Figure 5, aiming to provide a user-friendly experience for clinicians, patients and researchers.

**Fig. 5.**
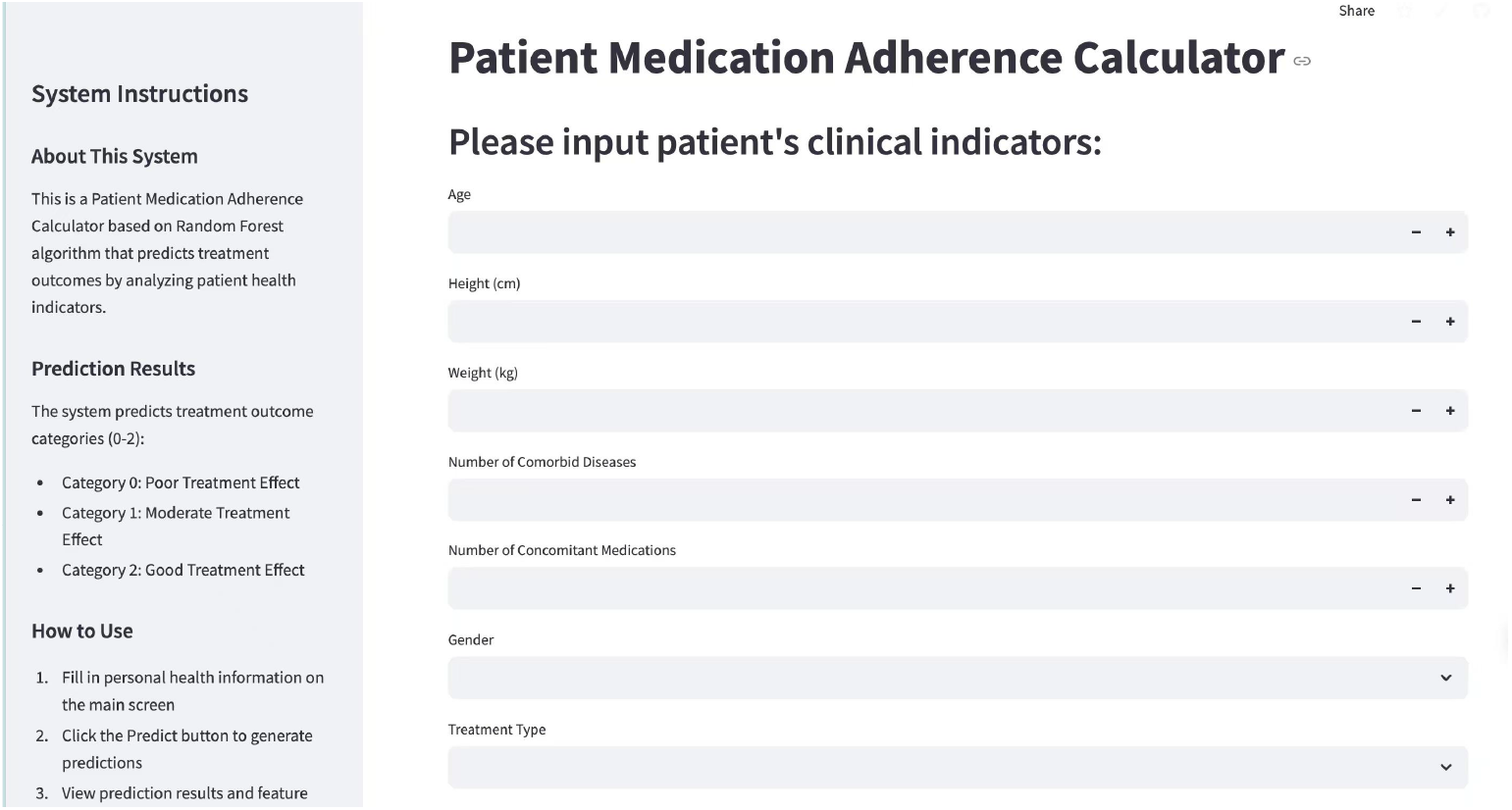
The screenshot of the web-based tool.

## Discussion

This study introduces the Random Forest Classifier (RFC) model[9]as a predictive tool for assessing medication adherence in patients using drugs, demonstrating superior performance over competing models across multiple metrics such as AUC, accuracy, precision, recall, F1 score, and kappa coefficient[10]. The model’s robustness in parsing complex medical data reveals its potential for accurate medication adherence risk predictions[11].

Unlike traditional assessment tools, which often rely on clinical judgment[12], the RFC model integrates and analyzes a broad range of clinical and laboratory data, automating the analysis process[13]. This circumvents the need for manual scoring by clinicians, facilitating quicker and more objective assessments[14]. The development of a web-based calculator further enhances the model’s clinical utility, providing healthcare professionals with a readily accessible tool for real-time medication adherence assessment[15].

This predictive model can assist the general population and potential drug users in prospectively assessing their own ability for drug management. By intelligently analyzing their daily medication behaviors and life routines, it can accurately identify potential tendencies of poor medication compliance. When the assessment results indicate a risk of insufficient compliance, users can promptly notice the weak links in their health management and actively optimize their behavior patterns through personalized methods such as setting medication reminders and integrating medication taking with fixed daily behaviors. This mechanism not only transforms abstract health management into concrete visual feedback but also forms a virtuous cycle through continuous behavioral guidance, helping users cultivate rigorous medication awareness subconsciously and establish scientific and standardized medication habits in the early stages of disease prevention or treatment.

This research contributes to the literature on machine learning (ML) applications in refining medication adherence assessment strategies[16], expanding upon previous studies[17]. It acknowledges the multifactorial nature of medication non-adherence, arising from various factors such as patient demographics, comorbidities, medication regimens, and psychosocial factors[18], which manifest the need for comprehensive approaches to understanding and managing this condition[19]. The RFC model marks a notable advancement in applying ML to patient care, offering a nuanced and efficient method for identifying at-risk individuals[20].

The relationship between demographic factors (such as age and gender) and medication adherence is well-documented[21]. Our study highlights that higher age and male gender are associated with poorer medication adherence, which is consistent with previous findings[22]. Additionally, the presence of multiple comorbid diseases and higher BMI also correlate with non-adherence[23], suggesting that these factors may complicate medication management and patient engagement[24].

The impact of medication regimen complexity on adherence is another critical area explored in this study[25]. The findings indicate that a higher number of concomitant medications and polypharmacy are significant predictors of poor adherence[26]. This underscores the importance of optimizing medication regimens and minimizing unnecessary polypharmacy to improve patient outcomes[27].

Geographical location and educational attainment also play a role in medication adherence[28-29]. Patients from urban areas and those with higher educational levels tend to have better adherence, possibly due to improved access to healthcare resources and better understanding of treatment plans[30]. These findings suggest that targeted interventions may be needed for patients in rural areas or those with lower educational backgrounds[31].

Our study also highlights the importance of regular follow-up and the presence of a history of stroke in predicting medication adherence[32]. Regular follow-up visits provide opportunities for healthcare providers to reinforce the importance of adherence and address any barriers or side effects[33]. The association between a history of stroke and better adherence may reflect heightened awareness of the risks associated with non-adherence among these patients[34].

The utilization of SHAP (SHapley Additive exPlanations) for model interpretation further elucidates the contributions of specific variables to medication adherence risk, enhancing the model’s clinical relevance[35]. The SHAP values provide insights into how each feature influences the model’s predictions, allowing clinicians to identify key factors driving non-adherence in individual patients[36].

Most existing studies focus on compliance assessment based on patients’ current clinical characteristics or historical behaviors. Although such methods can identify the current situation, they are difficult to achieve early risk warning and dynamic intervention. The innovation of this study lies in integrating multi-dimensional patient data (such as demographic characteristics, comorbidities, medication complexity, etc.) to construct a machine learning model with forward-looking predictive capabilities.

This model can not only assess the compliance status of patients in real time, but also predict their potential future compliance risk levels (such as low, medium, and high) based on individual characteristics. This predictive ability makes it possible for clinical intervention to shift from traditional passive response to active prevention.

For example, for patients predicted to be at high risk, personalized intervention measures (such as simplifying medication regimens, strengthening medication reminders, or providing psychological support) can be implemented in advance to effectively block the risk chain before compliance deteriorates. In addition, the dynamic prediction feature of the model can be combined with the patient’s real-time updated clinical data to achieve dynamic adjustment of risk stratification and intervention strategies, providing technical support for full-cycle health management in the era of precision medicine. Compared with static assessment tools, this study opens up a new path for improving the treatment outcomes of long-term medication patients through the combination of predictive modeling and early intervention.

Despite its promising results, our study has some limitations. The data were collected from a single center, which may introduce biases and limit the generalizability of the findings . Future research should focus on expanding the model’s generalizability through multi-center studies . Additionally, the exclusion of variables with excessive missing data points might have affected the model’s performance . Future studies should aim to improve data collection methods to minimize missing data . Furthermore, the model’s performance could be further enhanced by incorporating additional biomarkers and psychosocial factors that may influence medication adherence .

In conclusion, this study demonstrates the potential of advanced ML techniques in transforming the approach to medication adherence assessment within the medical field . The RFC model outperformed existing assessment tools and other ML models in accuracy and efficiency . Deployed as a web-based application, the model offers a practical tool for healthcare professionals, facilitating early identification and management of medication non-adherence .Furthermore, this model can also assist users in cultivating a rigorous medication awareness unconsciously, and establish scientific and standardized medication habits at the early stage of disease prevention or treatment. Future research should focus on expanding the model’s generalizability and integrating additional factors to refine its predictive capability .

## Conclusions

This study introduces a novel multi-classification machine learning (ML)-based model, employing the Random Forest Classifier (RFC) algorithm, for the assessment of medication adherence in patients using drugs. Through rigorous analysis of multidimensional patient data, including clinical information, laboratory test results, and medication regimens, we identified key variables substantially associated with medication adherence risk. The RFC model outperformed existing assessment tools and other ML models in accuracy and efficiency.

The utilization of SHAP for model interpretation further elucidates the contributions of specific variables to medication adherence risk, enhancing the model’s clinical relevance. Deployed as a web-based application, the model offers a practical tool for healthcare professionals, facilitating early identification and management of medication non-adherence in patients using drugs.

Despite its promising results, future research should focus on expanding the model’s generalizability through integrating other indicators to refine its predictive capability. This study displays the potential of advanced ML techniques in transforming the approach to medication adherence assessment within the medical field, particularly for patients with complex chronic conditions requiring long-term therapy.

Data availability Data presented in this study were obtained from patients admitted to Nanjing Drum Tower Hospital who consented to participate in the study. Owing to data protection rules, we are not allowed to share personal-level data. The datasets used and/or analysed during the current study are available from the corresponding author on reasonable request.

## Data Availability

All data produced in the present study are available upon reasonable request to the authors.

